# Extracellular Matrix Abnormalities in the Hippocampus of Subjects with Substance Use Disorder

**DOI:** 10.1101/2023.09.07.23295222

**Authors:** Jake Valeri, Charlotte Stiplosek, Sinead M. O’Donovan, David Sinclair, Kathleen Grant, Ratna Bollavarapu, Donna M. Platt, Craig A. Stockmeier, Barbara Gisabella, Harry Pantazopoulos

**Affiliations:** Department of Psychiatry and Human Behavior, University of Mississippi Medical Center, Jackson, MS; Program in Neuroscience, University of Mississippi Medical Center, Jackson, MS; Department of Neuroscience, University of Toledo, Toledo, OH; Oregon Primate Research Center, Hillsboro, OR

## Abstract

Contextual triggers are significant factors contributing to relapse in substance use disorders (SUD). Emerging evidence points to a critical role of extracellular matrix (ECM) molecules as mediators of reward memories. Chondroitin sulfate proteoglycans (CSPGs) are a subset of ECM molecules that form perineuronal nets (PNN) around inhibitory neurons. PNNs restrict synaptic connections and help maintain synapses. Rodent models suggest that modulation of PNNs may strengthen contextual reward memories in SUD. However, there is currently a lack of information regarding PNNs in the hippocampus of people with SUD as well as how comorbidity with major depressive disorder (MDD) may affect PNNs. We used postmortem hippocampal tissues from cohorts of human and nonhuman primates with or without chronic alcohol use to test the hypothesis that PNNs are increased in subjects with SUD. We used histochemical labeling and quantitative microscopy to examine PNNs, and qRT-PCR to examine gene expression for ECM molecules, synaptic markers and related markers. We identified increased densities of PNNs and CSPG-labeled glial cells in SUD, coinciding with decreased expression of the ECM protease matrix metalloproteinase 9 (*Mmp9*), and increased expression for the excitatory synaptic marker vesicle associated membrane protein 2 (*Vamp2*). Similar increases in PNNs were observed in monkeys with chronic alcohol self-administration. Subjects with MDD displayed changes opposite to SUD, and subjects with SUD and comorbid MDD had minimal changes in any of the outcome measures examined. Our findings demonstrate that PNNs are increased in SUD, possibly contributing to stabilizing contextual reward memories as suggested by preclinical studies. Our results also point to a previously unsuspected role for CSPG expression in glial cells in SUD. Evidence for increased hippocampal PNNs in SUD suggests that targeting PNNs to weaken contextual reward memories is a promising therapeutic approach for SUD, however comorbidity with MDD is a significant consideration.

## INTRODUCTION

Substance use disorders (SUD) are a group of psychiatric disorders with substantially increased risk of mortality and severe socioeconomic burden affecting approximately 7 % of people in the United States annually ^1, 2^. SUDs impose a substantial burden of morbidity and mortality, exacerbated by persistently high rates of relapse and treatment failure ^3^. Contextual cues associated with drug use are a critical factor contributing to relapse in individuals with SUD and highlight the strength of memory circuits associated with reward processing ^3-7^. A growing number of studies indicate a critical role of extracellular matrix molecules (ECM) in the regulation of reward memories ^8-17^ (for reviews see ^18, 19^). Recent gene expression profiling studies demonstrate altered expression of pathways involved in ECM regulation in the dorsolateral prefrontal cortex and nucleus accumbens of subjects with opioid use disorder (OUD) ^12, 20^ as well as in preclinical models of OUD ^20^.

Preclinical studies of cocaine, alcohol or opioid use demonstrate that alteration of specialized ECM structures called perineuronal nets (PNN) represent a key feature underlying reward memory in SUDs ^8, 9, 17, 21-24^. PNNs are ECM structures that envelop populations of fast-firing inhibitory neurons, stabilizing synapses on those neurons and restricting synaptic plasticity ^16, 22, 25, 26^. During acquisition of drug memories, endogenous proteases including matrix metalloproteases (MMPs) degrade PNN components to allow for formation of new synapses associated with reward memories. ^11, 27-30^. Chronic drug use resulting in prolonged re-activation of memory circuits results in enhancement of PNN components in the prefrontal cortex and insular cortex, stronger than at baseline, which stabilizes reward-associated synapses ^9, 23, 24, 31-33^ and may contribute to context-induced relapse ^11, 27-30^. Evidence that intracerebral injection of a pharmacological MMP inhibitor significantly weakens cocaine reward memories provides further support for this hypothesis ^10^. The hippocampus is at the center of neurocircuitry involved in reward memory processing ^34^. Specifically, accumulating evidence suggests that hippocampal sector CA1 is involved in integrating dopamine reward signaling with contextual memories and in regulating reward expectation and extinction ^35-37^. CA1 neurons receive dopaminergic projections from the ventral tegmental area and encode reward-associated spatial memories ^37, 38^. CA1 spatial maps are modified by reward expectation and by inhibitory control of excitatory neurons encoding these maps ^37, 38^. Inhibitory neurons in CA1 are often surround by PNNs, and alterations of hippocampal PNNs impact several neurotransmitter systems associated with reward processing. For example, PNN degradation alters firing rate of inhibitory parvalbumin (PVB) neurons ^39^, as well as NMDAR/AMPAR trafficking ^40^ and hippocampal dopamine transmission ^41^.

Despite this evidence, there is a lack of information regarding PNNs in the hippocampus of subjects with SUD, limiting the translatability of preclinical findings and the development of ECM-based therapies for context-induced relapse. Furthermore, subjects with SUD often have comorbid substance use and a high degree of comorbidity with major depressive disorder (MDD)^42^. These features are challenging to capture in preclinical models, potentially limiting translatability. We used a cohort of human postmortem samples from subjects with SUD with or without comorbid MDD to test the hypothesis that numerical density of PNNs is increased in sector CA1 of subjects with SUD, accompanied by altered expression of genes related to ECM degradation and biosynthesis, synaptic regulation, and PVB expression. This cohort includes retrospective clinical assessments, toxicology reports, history of substance use and medication history, allowing for testing of several of the features inherent in the population with SUD. Subjects with comorbid SUD and MDD, and subjects with MDD without SUD were included to examine the potential effects of comorbid MDD on ECM pathology in SUD. Furthermore, we used a collection of postmortem hippocampus samples from rhesus monkeys with or without chronic alcohol self-administration to examine the direct cause and effect on PNNs of the most common drug of abuse present across all our subjects with SUD.

## MATERIALS AND METHODS

### Postmortem brain samples

#### Human subjects for brightfield microscopy and RNA studies

Fresh frozen blocks containing the hippocampus from subjects with SUD (n=20), SUD and comorbid MDD (n=24), MDD (n=20), and psychiatrically normal controls (n=20) were provided by the UMMC Postmortem Brain Core (Table 1, Cohort A). Details are described in our previous report ^43^ and in the Supplemental Materials.

**Table 1.** Basic cohort demographic information for both human and rhesus macaque subjects. Abbreviations: SUD, substance use disorder; MDD, major depressive disorder; PFA: Paraformaldehyde; PMI, postmortem interval, BAC, blood alcohol concentration, 12-month average. Values represent mean ± SEM.

| Cohort A: Human subjects for SUD, SUD/MDD, and MDD analyses |  |  |  |  |
| --- | --- | --- | --- | --- |
|  | SUD<br>*n*=20 | SUD/MDD<br>*n*=24 | MDD<br>*n*=20 | Control<br>*n*=20 |
| Age, Years | 34.8±9.0 | 43.9±11.1 | 47±11.4 | 43.2±11.42 |
| Sex |  |  |  |  |
| Male | 17 | 16 | 14 | 13 |
| Female | 3 | 8 | 6 | 7 |
| Race |  |  |  |  |
| White | 13 | 20 | 17 | 11 |
| Black | 7 | 4 | 3 | 9 |
| Brain pH | 6.57±0.23 | 6.54±0.27 | 6.5±0.32 | 6.5±0.25 |
| PMI, Hours | 18.58±8.5 | 20.2±7.6 | 23.7±5.9 | 21.2±6.8 |
| Cohort B: PFA fixed samples from control subjects for labeling of synaptic markers and PVB neurons |  |  |  |  |
| Age | 43.0±25.2 |  |  |  |
| Sex | 2 Male, 1 Female |  |  |  |
| Hemisphere | 2 Right, 1 Left |  |  |  |
| Cohort C: Rhesus macaque subjects |  |  |  |  |
|  | Alcohol drinkers<br>*n*=7 |  | Control<br>*n*=5 |  |
| Age, Years | 7.20±0.04 |  | 6.90±0.49 |  |
| Sex | Male (*n*=7) |  | Male (*n*=5) |  |
| BAC, mg pct | 101.46±12.76 |  | NA |  |

#### Psychiatric control subjects for confocal imaging

Paraformaldehyde fixed free-floating samples containing the hippocampus were obtained from a separate cohort of three control subjects from the Harvard Brain Tissue Resource Center (Table 1, Cohort B), as fresh frozen tissue is not suitable for reliable immunohistochemical labeling for parvalbumin and synaptic markers. Two psychiatrists determined the absence of DSM-IV diagnoses based on the review of a questionnaire filled out by legal next of kin and a review of all available medical records. Control cases had sufficient information from legally-defined next of kin and medical records to rule out major medical, neurologic, and psychiatric conditions. All brains underwent a complete neuropathological exam and cases with histopathological abnormalities were excluded from this study.

#### Rhesus macaque subjects

Fresh frozen hippocampus samples from adult male rhesus macaques (*Macaca mulatta*) were obtained from the Monkey Alcohol Tissue Research Resource (MATRR) Cohort 5 (Table 1, Cohort C). Details regarding these subjects are available at www.matrr.com and described in the Supplemental Materials. Subjects were allowed to freely self-administer alcohol for 22 h daily for one year.

### Tissue collection and processing

#### Fresh frozen postmortem samples (Cohorts A&C)

Coronal serial sections (14 μm) were obtained from fresh frozen postmortem hippocampal blocks. Blocks were dissected and rapidly frozen in 2-methylbutane and on dry ice without fixation and kept in dry ice before permanent storage at -80 °C. Blocks were sectioned using a Leica CM3050S cryostat.

#### Paraformaldehyde fixed control subjects (Cohort B)

Tissue blocks containing the hippocampus were processed as described previously ^44^.

### Histochemical/immunocytochemical labeling, imaging, and quantification procedures

#### Brightfield microscopy

Fresh-frozen, slide-mounted medial hippocampal cryosections from human subjects and rhesus macaque subjects (3-5 sections per subject) were post-fixed for 30 minutes in 4% paraformaldehyde, followed by 1-hour incubation in 2% bovine serum albumin (BSA), and overnight incubation in biotinylated *Wisteria floribunda agglutinin* (WFA) lectin (1:500, catalog #B-1355, Vector Labs). WFA binds to non-sulfated N-acetyl-D-galactosamine residues on the terminal ends of CSPGs ^45^. The following day, tissue was incubated in streptavidin horseradish peroxidase (1:5000 μl, Zymed, San Francisco, CA) for four hours, followed by a 20-minute incubation in 3’3-diaminobenzidine with nickel sulfate hexahydrate and H_2_O_2_. Tissue was dehydrated through an ethanol and xylene series, followed by histological labeling with Methyl Green (catalog #H-3402, Vector Labs). Sections were coverslipped and coded for blinded quantitative analysis. All sections included in the study were processed simultaneously to avoid procedural differences. Quantification was performed using an Olympus BX61 microscope and WFA-labeled PNNs were distinguished from WFA+ glial cells based on morphology. Each anatomical subdivision of the hippocampus was traced for an area measurement using a 20X objective, and WFA-labeled elements were quantified on 40X magnification using Stereo-Investigator v11.

#### Immunofluorescence and Imaging

Immunofluorescence for VAMP2-PVB-WFA and for GFAP-WFA were conducted as described previously ^46, 47^ and in the Supplemental Materials.

### Quantitative polymerase chain reaction

qPCR was conducted as described previously ^43, 48^ and in the Supplemental Materials. The primers used are listed in Supplemental Table 21.

### Statistical Analysis

Numerical densities of microscopy measures were calculated as described in our previous studies ^49 46, 47^. The goal of our study was to test for differences in PNN densities in CA1 and hippocampal gene expression of ECM and synaptic markers between diagnosis groups. Analysis of WFA-glial densities was focused on the DG and CA4 areas where these cells are primarily distributed in the hippocampus in our current study and our previous report ^49^. Diagnosis groups with comorbid SUD/MDD and MDD were included to test for potential effects of comorbid depression on findings in SUD. Exploratory analysis was conducted on microscopy measures of PNNs and WFA-glia on additional hippocampal sectors. Differences between groups relative to the main outcome measures were assessed for statistical significance using stepwise linear regression analysis of covariance (ANCOVA) using JMP Pro v16.1.0 (SAS Institute Inc., Cary, NC). Logarithmic transformation was applied to values when not normally distributed. Potential confounding variables (Table 1 and Supplementary Tables 5-20) were tested systematically for their effects on main outcome measures and included in the model if they significantly improved goodness-of-fit. Covariates found to significantly affect outcome measures are reported. Subjects with SUD, subjects with SUD/MDD, and subjects with MDD were first compared separately with psychiatrically-normal controls. Subsequently, the four groups were considered together to test for differences between diagnostic groups and Bonferroni correction applied for multiple comparisons.

## RESULTS

### Altered PNN and WFA-glia densities in subjects with SUD, MDD, and comorbid SUD/MDD

Numerical densities of PNNs were significantly increased in CA1 stratum oriens (SO) of subjects with SUD (p < 0.007; adjusted for significant effects of PMI and cocaine history). (Figure 1A-E), where the majority of PNNs were distributed in sector CA1 (Control CA1 SO: 3.5 PNNs/mm^2^; Control CA1 SP: 0.97 PNNs/mm^2^). Further analysis of additional hippocampal sectors revealed increased numerical densities of PNNs in CA2 SO (p < 0.03; adjusted for significant effects of age), CA2 SP (p < 0.0001; adjusted for significant effects of cocaine history), CA3 SO (p <0.01; adjusted for significant effects of cocaine history), and CA4 (p < 0.007; adjusted for significant effects of cocaine history) in subjects with SUD (Figure 1G, Table S1). History of cocaine use, which was a significant covariate across hippocampal subregions, was associated with decreased PNNs in each of these regions (Figure S4D). No significant differences were found in the dentate gyrus or the CA1 and CA3 SP layers. In subjects with MDD, we observed significantly decreased numerical densities of PNNs in CA1 SO (p < 0.05; adjusted for significant effects of sex and calcium channel blockers) and CA1 SP (p < 0.02), increased PNNs in CA4 (p < 0.01; adjusted for significant effects of anxiety disorder diagnoses), and no significant differences in other regions of the hippocampus. The duration of depression positively correlated with PNN density in CA1 SO across subjects with SUD/MDD and MDD (Figure S1A). In subjects with comorbid SUD/MDD, we observed reduced numerical densities of PNNs in CA1 SP (p < 0.02; adjusted for significant effects of substance abuse history and duration of MDD, Figure S1B), increased densities of PNNs in CA2 SP (p < 0.01; adjusted for significant effects of SSRIs in the toxicology report and ZT time). No significant differences were observed in the other hippocampal subregions. Table S1 contains information for each hippocampus subregion, including p-values, F-ratios, adjusted least squares means, standard error, and significant covariates.

**Figure 1.**
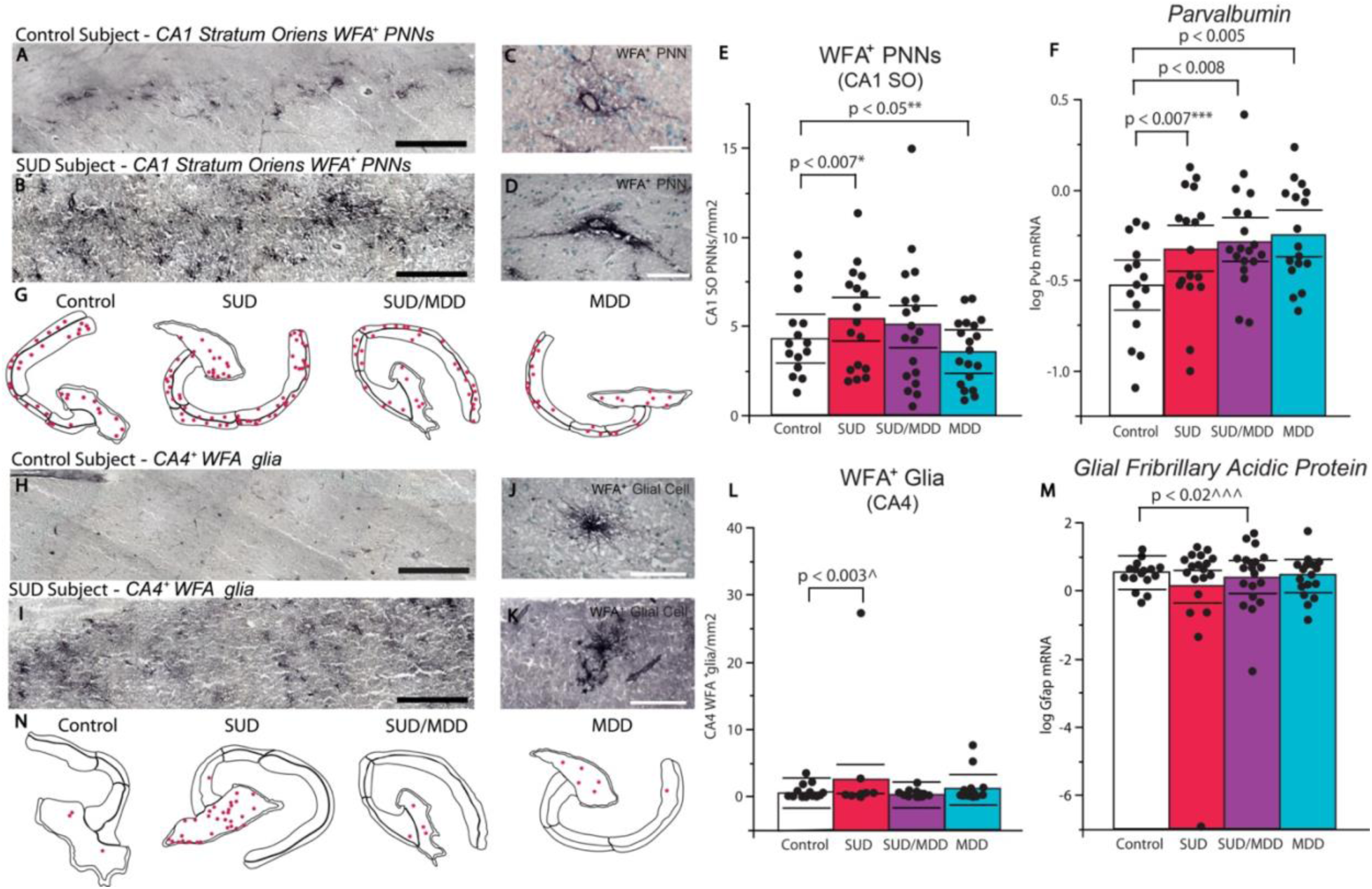
Altered PNNs in SUD and MDD. (A and B) Representative low-magnification photomicrographs of CA1 SO PNNs labeled by WFA, scalebars = 0.5 mm. (C and D) High-magnification photomicrographs of WFA-labeled CA1 SO PNNs, scalebars = 50 µm. (E) Diagnostic group comparisons of PNN densities in CA1 stratum oriens. Subjects with SUD had significantly increased densities of PNNs (p < 0.007; adjusted for significant effects of cocaine history and PMI) whereas subjects with MDD displayed significantly reduced PNN densities (p < 0.05; adjusted for significant effects of sex and calcium channel blockers). No significant effects were found in the comorbid SUD and MDD group. (F) Diagnostic group comparisons of normalized hippocampal *Pvb* gene expression, with a gradient increase between SUD (p < 0.007; adjusted for significant effects of cocaine in the toxicology report), SUD/MDD (p < 0.008; adjusted for significant effects of MDD duration), and MDD (p < 0.005). (G) Representative PNN quantification overlays of hippocampi from subjects within diagnostic groups. (H and I) Representative low-magnification photomicrographs of CA4 WFA-labeled glia, scalebars = 0.5 mm. (J and K) Representative high-magnification WFA-labeled glia, scalebars = 80 50 µm. (L) Numerical density measurements of CA4 WFA-labeled glial cells in subjects with SUD, MDD, and SUD/MDD. (M) Normalized gene expression of *Gfap* was significantly decreased in subjects with comorbid SUD and MDD (p < 0.02; adjusted for significant effects of antidepressant history), but unaffected in either isolated disorder. (N) Representative WFA+ glia quantification overlays of hippocampi from subjects within diagnostic groups. Error bars are mean ± SEM.

We observed WFA-labeled glial cells in addition to PNN labeling. WFA-glia in our current study were shown to co-express the astrocyte marker glial fibrillary acidic protein (GFAP) (Figure S5) and were morphologically similar to WFA-glia described our previous reports in the human medial temporal lobe ^46, 47^. WFA-glial cells were distributed primarily in the dentate gyrus and CA4 areas, with sparse expression in other hippocampal subregions (Figure 1H-K, N). In subjects with SUD, we observed significantly greater density of WFA-glia in CA4 (p < 0.002; adjusted for significant effect of cocaine history), and in the dentate gyrus (p < 0.02; adjusted for significant effect of cocaine history, Figure 1L). No other hippocampal subregions displayed statistically significant differences in subjects with SUD. In subjects with MDD, we observed significantly increased densities of WFA-glia in the dentate gyrus (p < 0.0001), CA3 SP (p < 0.0008; adjusted for a significant effect of antidepressants in last month of life), CA2 SP (p < 0.006; adjusted for a significant effect of antidepressants in last month of life), and CA2 SO (p < 0.02). All other hippocampal subregions lacked differences in subjects with MDD. In subjects with comorbid SUD/MDD, we observed significantly decreased numerical densities of WFA-glia in the dentate gyrus (p < 0.02; adjusted for significant effects of alcohol in the toxicology report and calcium channel blockers), CA4 (p < 0.02; adjusted for a significant effect of alcohol in the toxicology report), CA3 SP (p < 0.002; adjusted for significant effects of alcohol in the toxicology report and race), CA2 SP (p < 0.004; adjusted for a significant effect of race), CA1 SP (p < 0.02; adjusted for significant effects of antipsychotics and alcohol in the toxicology report), and CA1 SO (p < 0.02; adjusted for a significant effect of race) when compared to psychiatric control subjects. Table S2 contains information for each hippocampus subregion. No differences in hippocampal area measurements were detected between control subjects and any of the diagnosis groups (Supplementary Table S4).

### Gene expression of markers for cell populations associated with PNNs and WFA-glial cells

#### Parvalbumin

Neurons expressing parvalbumin (*Pvb*) mRNA represent the majority of neurons surrounded by PNNs in the hippocampus ^49-51^. Subjects with SUD had significantly greater gene expression of *Pvb* compared to control subjects (Fig. 1F, Table S3; p <0.007; adjusted for a significant effect of cocaine history). Subjects in the SUD group with cocaine in the blood at death had significantly lower *Pvb* gene expression compared to subjects with SUD without cocaine in the blood at death (Supplemental Figure S4B). *Pvb* mRNA was also significantly greater in subjects with comorbid SUD/MDD (p < 0.008), and subjects with MDD (p < 0.005) compared to control subjects (Fig. 1F, Table S3).

#### Glial fibrillary acidic protein

The astrocyte marker glial fibrillary acidic protein (Gfap) colocalized with astrocytes labeled by WFA (Figure S5), which is in line with previous reports from human postmortem studies ^46, 52^. *Gfap* gene expression was significantly lower in the hippocampus of subjects with comorbid SUD/MDD (p < 0.02; adjusted for significant effects of SSRIs in last month of life which increases *Gfap*) (Fig. 1M, Table S3). No changes were detected in the SUD or MDD groups.

### Altered expression of synaptic markers in the hippocampus of subjects with SUD

#### Synaptobrevin and Synapsin 1

Synaptobrevin (VAMP2) is an excitatory synaptic marker that contributes to the SNARE complex, allowing for fusion of vesicles to the cell membrane to facilitate neurotransmitter exocytosis and receptor trafficking ^53, 54^. We examined VAMP2 as a step towards altered synaptic regulation in the hippocampus of subjects with SUD as suggested by extensive literature indicating altered synaptic regulation by chronic exposure to substances of abuse (for review see ^55^). We first tested whether VAMP2 protein is located on PVB neurons and in PNNs, and observed VAMP2 protein on PVB neurons ensheathed by PNNs and enmeshed within the PNN (Figure 2B-E). In subjects with SUD, *Vamp2* mRNA was significantly increased (p < 0.04; adjusted for a significant effect of sleep quality). An increase in *Vamp2* transcription was also observed in subjects with MDD (p < 0.005; with significant effects of age, ZT time, and sleep disturbance). No changes in *Vamp2* expression were detected in subjects with comorbid SUD/MDD. We conducted similar analysis for the inhibitory presynaptic marker, synapsin 1 (*Syn1*). We observed spatial overlap of SYN1 protein with PVB interneurons coated by PNNs, including SYN1 within PNNs (Figure 2G-J). In contrast to Vamp2, no significant differences were observed for *Syn1* gene expression in any of the diagnostic groups (Figure 2F).

**Figure 2.**
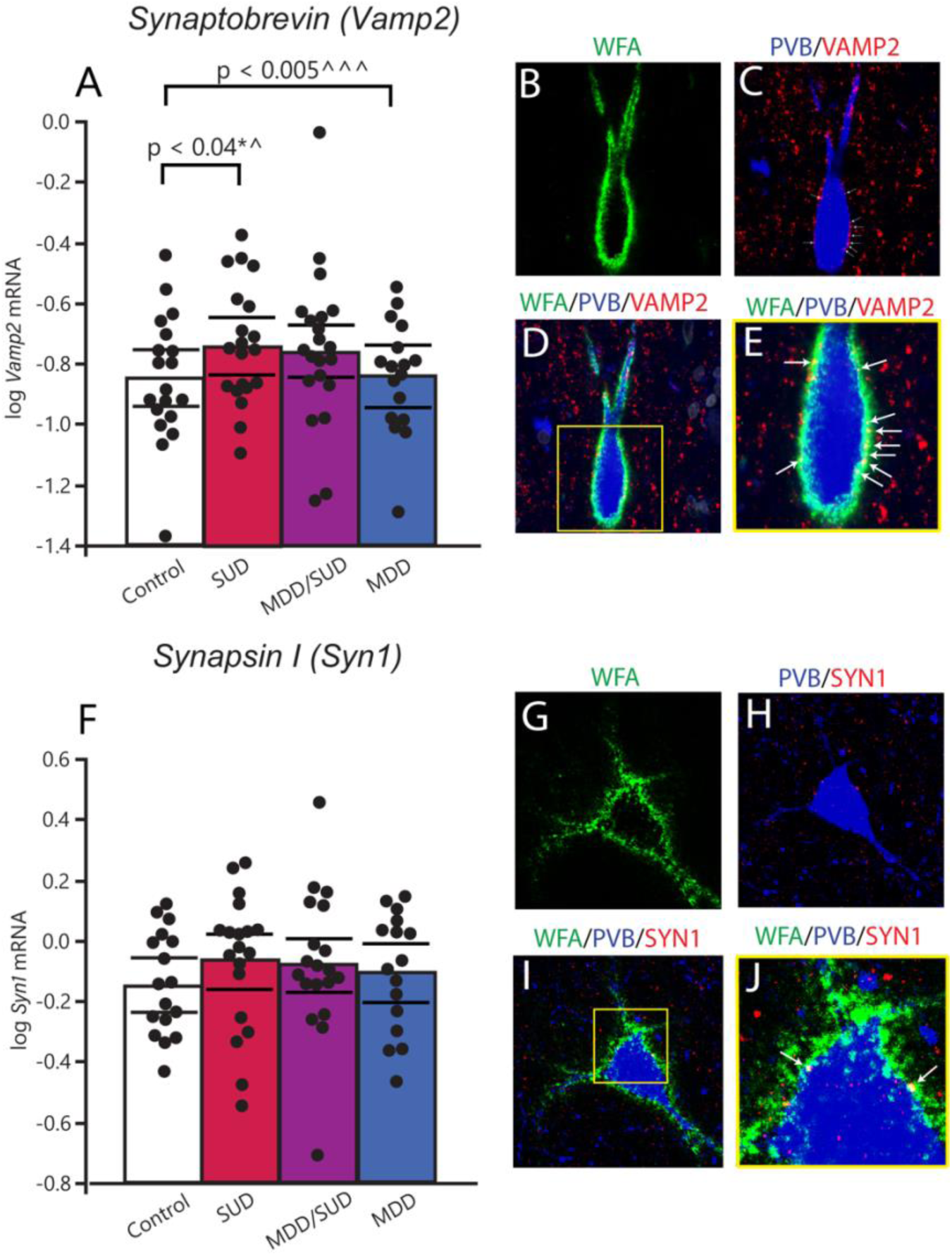
Gene expression of the excitatory synaptic marker Vamp2 is altered in SUD and MDD. (A) Gene expression of the presynaptic vesicle marker *Vamp2* is increased in both SUD and MDD, but not in the comorbid condition. (B-E) Multiplex immunofluorescent imaging of VAMP2 immunoreactive puncta within a PNN and on the surface of a PVB interneuron. (B) WFA-labeled PNN (green), (C) PVB interneuron (blue) and VAMP2 synaptic puncta (red) and spatial overlap (yellow) (arrows indicate overlap). (D) Composite z-projection of WFA, PVB, and VAMP2. (E) Zoomed inset of 1D, with arrows denoting spatial overlap of VAMP2 with WFA (yellow puncta). (F) There were no significant changes in transcription of the synaptic marker *Syn1* in any of the diagnostic groups. (G-J) Multiplex immunofluorescent imaging depicting SYN1 labeling within a PNN surrounding a PVB interneuron. (G) WFA-labeled PNN (green), (H) PVB neuron (blue) and SYN1 (red), (I) composite z-projection of all three channels. (J) Zoomed inset depicts spatial overlap with of SYN1 puncta with WFA labeling (arrows to yellow puncta). All scalebars are 10 µm. Error bars are mean ± SEM.

### Altered expression of ECM molecules in the hippocampus of subjects with SUD

#### Matrix metalloproteinase 9 (Mmp9) and Cathepsin S (Ctss)

Significantly decreased expression of *Mmp9* was observed in subjects with SUD (Figure 3A: p < 0.04). Furthermore, *Ctss* mRNA was significantly decreased in subjects with SUD (Figure 3B, S4: p < 0.03; with significant effects of cocaine in the toxicology report), and increased in subjects with MDD (p < 0.04; adjusted for significant effects of alcohol use severity and ZT time). No significant changes were observed in subjects with comorbid SUD/MDD (Figure 3A-B).

**Figure 3.**
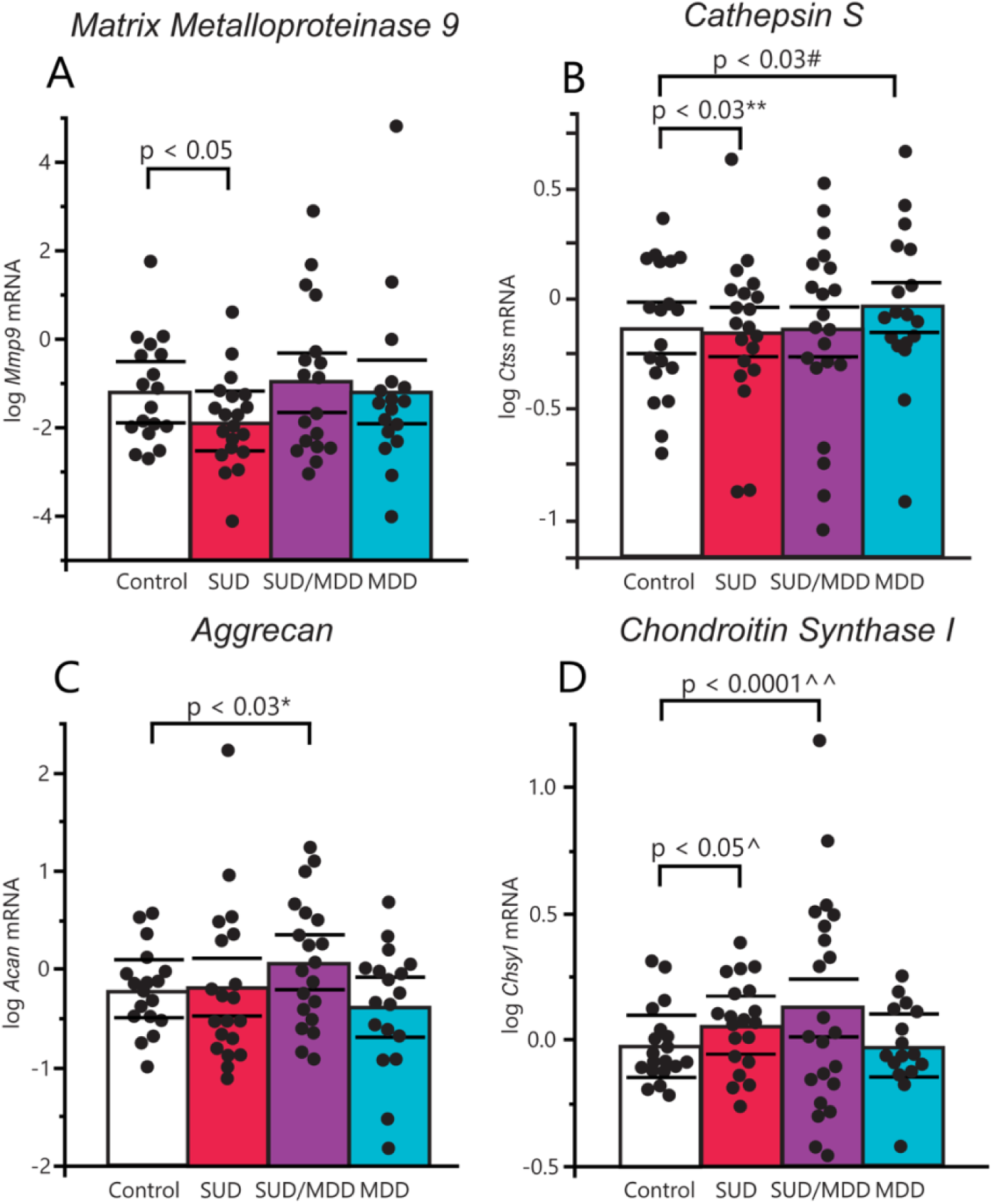
Gene expression of ECM biosynthetic and regulatory molecules in SUD and MDD. (A) Gene expression of the ECM proteolytic molecule *Mmp9* was significantly decreased in subjects with SUD compared to controls (p < 0.05), but no significant differences were detected in the other two diagnostic groups. (B) Analysis of covariance uncovered significantly decreased mRNA levels of cathepsin S in subjects with SUD (p < 0.03; adjusted for significant effects of cocaine in the toxicology report). Subjects with MDD had significantly increased expression of cathepsin S compared to controls (p < 0.03; adjusted for significant effects of alcohol history and ZT time of death). (C) Gene expression of the CSPG aggrecan was significantly increased in subjects with comorbid SUD and MDD (p < 0.03) but was unaffected in either condition separately. (D) Gene expression of the CSPG biosynthetic molecule *Chsy1* was significantly increased in subjects with SUD (p < 0.05; adjusted for significant effects of tissue pH) and in subjects with comorbid SUD and MDD (p < 0.0001; adjusted for significant effects of tissue pH and duration of AUD), while patients with MDD did not meet significance in any direction. Error bars are mean ± SEM.

#### Aggrecan and Chondroitin synthase 1

In subjects with SUD/MDD, we observed significantly increased aggrecan (*Acan)* (Figure 3C: p < 0.03), as well as chondroitin synthase 1 (*Chsy1)* mRNA (Figure 3D, S3B: p < 0.0001; with significant effects of duration of AUD and tissue pH). In subjects with SUD, a significant increase in *Chsy1* expression was detected (Figure 3C, S3: p < 0.04; with a significant effect of tissue pH), and no significant difference was observed for *Acan* (Figure 3D). No changes were observed in subjects with MDD for either *Acan* or *Chsy1* (Figure 3C&D).

### Increased numerical densities of perineuronal nets in the hippocampus of monkeys with chronic alcohol self-administration

We observed a significant increase of PNNs in the combined hippocampal subfields in rhesus monkeys with chronic alcohol self-administration (p < 0.03). Similar increases were observed in CA1 SO (Figure 4E; p < 0.04), CA3 SO (Figure 4E; p < 0.03), and CA3 SP (Figure 4E; p < 0.03). We also detected significantly increased numerical densities of WFA-labeled glia in the dentate gyrus granule cell layer (Fig. 4F; p < 0.04).

**Figure 4.**
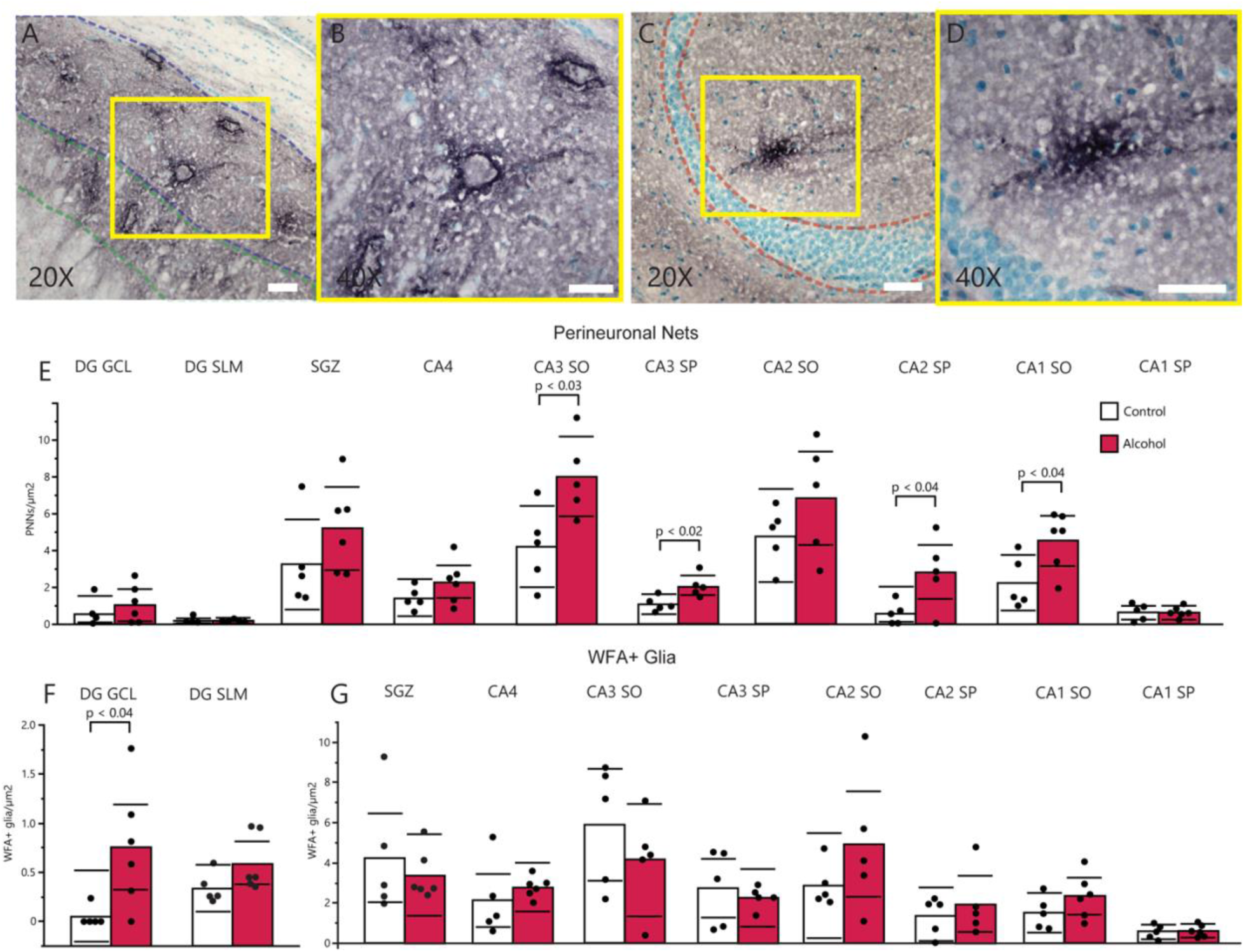
Chronic alcohol self-administration increases PNN densities throughout the hippocampus of rhesus monkeys. Abbreviations: DG GCL, granule cell layer of the dentate gyrus; SLM, stratum lacunosum moleculare; SGZ, subgranular zone; CA, cornu ammonis; SO, stratum oriens; SP, stratum pyramidale. (A) Representative microscopy of PNNs labeled with WFA in CA1 SO, scalebar = 50 µm. (B) Inset of Figure 4A at high-magnification, scalebar = 30 µm. (C) Representative microscopy of a WFA-labeled glial cell proximal to the DG GCL, scalebar = 50 µm. (D) Inset of Figure 4C at 40× magnification, scalebar = 50 µm. (E) Comparisons of PNN densities throughout the hippocampus between control (white bars) and monkeys with chronic alcohol self-administration (red bars). (F) Comparison of WFA-glia densities in the DG GCL and DG SLM between control and monkeys with chronic alcohol self-administration. (G) Comparison of WFA-glia densities in the SGZ and CA nuclei of the hippocampus between control and monkeys with chronic alcohol self-administration.

**Figure 5.**
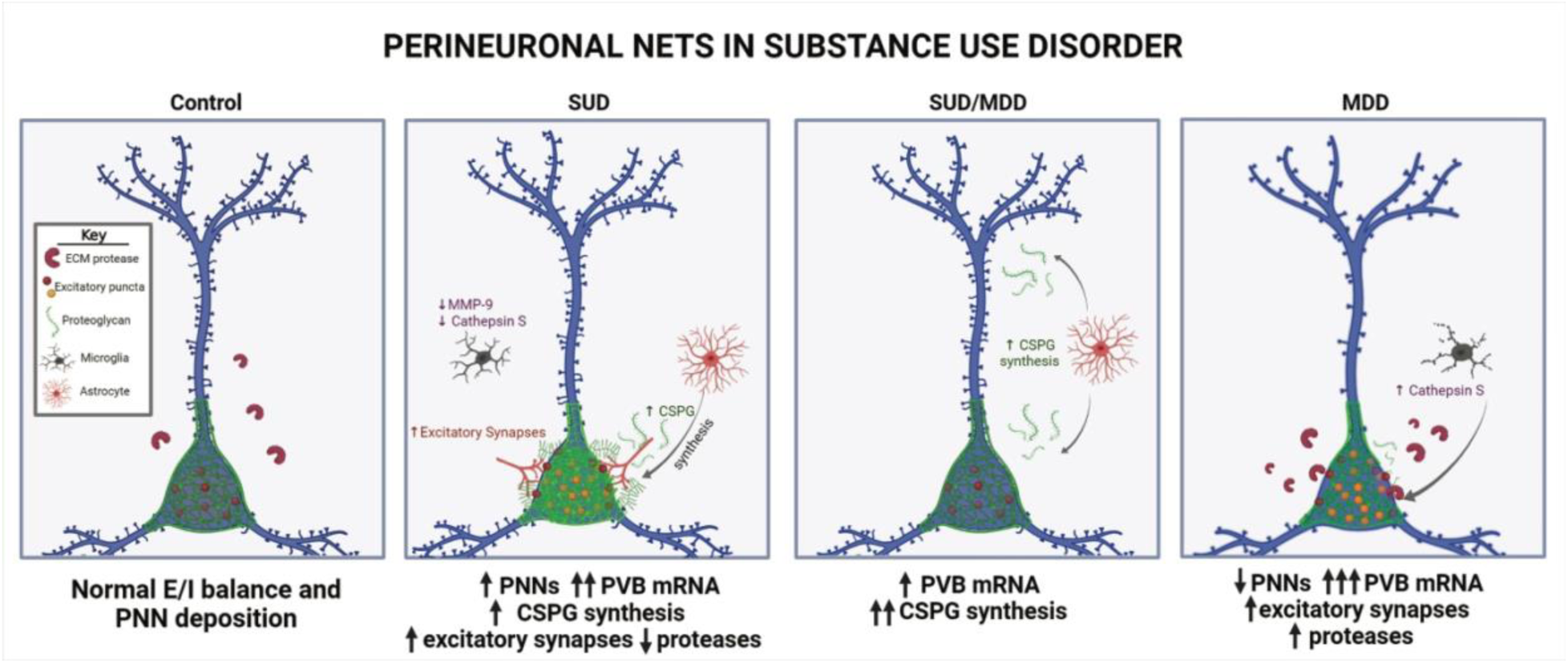
Summary diagram of proposed ECM and synaptic alterations in the hippocampus of subjects with SUD with or without comorbid MDD. Panels providing visual depictions of pathological observations from the present study. Numbers of arrows describe the intensity of changes compared to the other diagnostic groups, and a key is provided within the control panel to characterize the graphics used within the figure. Created with BioRender.com.

## DISCUSSION

Our data represent, to our knowledge, the first evidence for increased PNN densities in subjects with SUD and rhesus macaques with chronic alcohol self-administration. These findings link over a decade of evidence from preclinical models demonstrating increased PNNs in animals with chronic drug use ^9, 17, 23, 24, 32, 56-58^ with evidence in subjects with SUD. Together with the observed increased expression of the synaptic marker *Vamp2* and the CSPG synthesis molecule *Chsy1*, this suggests that enhanced PNN composition may aid in stabilizing contextual reward-associated synapses in the CA1 hippocampus of individuals with SUD. CA1 hippocampal neurons receive dopaminergic projections from the ventral tegmental area and encode reward-related spatial memories ^37, 38^. These CA1 spatial maps are modulated by local inhibitory control of excitatory neurons that encode the spatial map ^37, 38^. Speculatively, increased PNNs in CA1 of subjects with SUD may contribute to selective inhibitory control of excitatory neurons to sustain spatial maps involved in drug reward memories. Contextual memory processing may differ between drugs of abuse, and preclinical studies report decreases or no change in PNN densities with some drug classes including chronic nicotine, cocaine or opioid exposure ^17, 21, 31^. Our findings represent hippocampal alterations in subjects with polysubstance use which consists of a significant population of people seeking treatment for SUDs. Similar findings in nonhuman primates with chronic alcohol use together with increased PNNs in preclinical models of cocaine suggests that increased PNN densities may be a shared feature in people with polysubstance use disorder which often includes alcohol use disorder ^9, 24, 32, 59, 60^. Furthermore, decreased expression of ECM proteolytic molecules indicates that changes in PNN densities are accompanied by decreased ECM remodeling in chronic SUD. Increased expression of parvalbumin, expressed by neurons that PNNs typically encapsulate, suggests increased feedforward inhibition of local excitatory pyramidal neurons by parvalbumin that may enhance coherence of drug-associated neuronal ensembles. In contrast, decreased densities of PNNs in subjects with MDD represent the first evidence for altered PNNs in the hippocampus of subjects with MDD. Taken together, our findings support the hypothesis that increased PNNs may stabilize contextual reward memories in subjects with SUD and thus may represent therapeutic targets for context-induced relapse. Furthermore, decreased densities of PNNs in subjects with MDD and lack of significant differences in subjects with comorbid SUD/MDD highlight the importance of considering comorbid conditions for the neuropathology of SUD.

### PNNs and ECM molecules in the Hippocampus of Subjects with Substance Use Disorder

Our findings support an increasing number of preclinical studies demonstrating that PNNs stabilize reward memories ^8, 9, 16, 17, 22-24, 33^. Furthermore, PNNs are involved in a wide range of processes that can impact context-induced relapse including stabilization of synapses, receptor trafficking, and regulation of interneuron activity ^25, 26, 39, 40, 61-67^. Previous studies have implicated each of these processes in SUD. For example, chronic substance use has been shown to elicit downstream alteration of synapses and neurotransmitters including changes in dopamine 2 receptors (D2R) ^68^ as well as glutamatergic and GABAergic systems ^69, 70^. SUDs are chronically relapsing disorders which rely heavily on memory salience (i.e., cue responsiveness). PNNs contribute to synaptic homeostasis through the stabilization of synapses and PVB interneuron-mediated excitatory/inhibitory balance ^71^. Thus, PNNs may stabilize synapses involved in drug reward processes that contribute to context-induced relapse ^18^.

The association of PVB interneurons with PNNs has been demonstrated in several brain areas, including the hippocampus ^49, 72-74^. Hippocampal PVB neurons are predominantly located in the SO layers of the CA regions. PVB neurons are involved in modulating glutamatergic neurons and hippocampal output to brain regions involved in reward processing ^75, 76^. Increased PVB expression is positively correlated with PNN labeling ^77^ and removal of PNNs with chondroitinase ABC or antidepressants decreases PVB interneuron activity ^78^. Increased parvalbumin (PVB) expression in SUD may induce feedforward inhibition of local excitatory neurons that enhances coherence of drug-specific neuronal ensembles ^79^. Enhanced PNN composition on PVB neurons may also contribute to increased PVB expression and downstream consequences on excitatory/inhibitory balance which can result in altered activity of reward circuits involved in reward prediction and impulsivity ^80, 81^. Importantly, *Pvb* expression was significantly decreased in subjects with cocaine in their toxicology reports (Supplementary Figure 1G), suggesting that cocaine temporarily blunts PVB expression to disinhibit excitatory neurons^57^. Furthermore, decreased ECM proteolytic gene expression (*Mmp9* and *Ctss*) in subjects with SUD is in line with increased PNN densities in these subjects and with preclinical models of hippocampal MMP9 expression in alcohol use ^82^, potentially suggesting hypofunction of PNN proteolytic systems in SUD ^83^. In our ANCOVA models, subjects with SUD who had cocaine in their toxicology reports had greater expression of *Ctss* (Supplementary Figure 1H). Speculatively, increased *Ctss* and decreased *Pvb* in subjects with cocaine in the toxicology report may indicate that cocaine temporally activates *Ctss* to temporarily degrade PNNs and decrease in *Pvb* expression.

There is extensive evidence demonstrating a central role of the hippocampus for episodic memory processing ^84^. Furthermore, the hippocampus has bi-directional connections with the amygdala that contribute to assigning emotional valence to contextual memories, which is crucial in the long-term retention of drug memories ^85^. Hippocampal connections with the ventral tegmental area (VTA) are involved in regulating memory salience and reward predictions. Hippocampal glutamatergic fibers from CA1 travel through the subiculum to basal forebrain structures, and synapse onto VTA neurons which feed back to the hippocampus to release dopamine ^80, 81^. Thus, increased PNNs in the hippocampus of subjects with SUD can impact this system and contribute to several aspects of reward memory circuits.

Our studies represent first description of increased numerical densities of WFA-glial cells in individuals with SUD. Despite previous evidence that WFA-glia colocalize with the astrocytic marker GFAP ^46, 47^, we did not detect increased GFAP expression in subjects with SUD, suggesting that increases in WFA-glia do not reflect a broader astrogliosis process. Increased WFA-glia in the hippocampus may indicate increased CSPG production by glia contributing to increased PNNs in SUD.

We used a proximal specie, *Macaca mulatta*, with or without chronic alcohol use to test for specific effects of the most common drug in our cohort without the covariates inherent to cross-sectional human studies. Increased PNN densities and WFA-glia in rhesus monkeys with chronic alcohol use provides support for our findings in human subjects. Furthermore, the ability of chronic alcohol use alone to create these changes is intriguing and represents the first evidence for increased PNNs in the hippocampus of non-human primates in response to any drug of abuse. The present findings also represent the first observation of WFA-labeled glia in the non-human primate brain; to this point, they have only been reported in human subjects ^46, 49, 73^. These WFA-glial cell increases may represent enhanced CSPG synthesis by astrocytes.

### PNNs and ECM molecules in the Hippocampus of Subjects with Comorbid Substance Use Disorder and Major Depressive Disorder

Subjects with comorbid SUD and MDD represent a significant population of people with SUD and a bi-directional disorder where dysphoria can fuel the inclination to self-medicate. Alternatively, chronic drug use can contribute to mood disturbances ^86^. Approximately one third of patients with MDD also experience SUD, and conversely a substantial proportion of individuals with SUD will experience MDD at some point during prognosis ^42, 87^. This bi-directional comorbidity contributes to a heightened risk of suicide and greater social and personal impairment ^88^. Despite this relationship, co-occurrence of MDD and SUD is not typically examined in postmortem brain studies. Our results suggest MDD comorbidity is an important factor in studies of SUD. Inclusion of subjects with MDD may either mask or amplify changes present in each disorder. Most of our outcome measures displayed opposing directionality between subjects with SUD and MDD. For example, density of CA1 SO PNNs was significantly increased in SUD, decreased in MDD, and unaltered in subjects with comorbid SUD/MDD. In comparison, a comorbid diagnosis of MDD appeared to significantly amplify the existing increased expression of *Chsy1* observed in subjects with SUD (Figure 3D).

Increased expression of *Chsy1* in SUD and subjects with SUD/MDD may suggest that increased biosynthesis of CSPGs by chronic drug use is enhanced by processes underlying MDD. Expression of the CSPG aggrecan, a major PNN component ^89^, was increased in subjects with comorbid SUD/MDD. This may represent alterations in PNN composition that are not detected by standard WFA labeling.

### PNNs and ECM molecules in the Hippocampus of Subjects with Major Depressive Disorder

A strength of our cohort was the ability to cross-examine the incidence of MDD comorbid with SUD as well as subjects with MDD only. While densities of PNNs were decreased in subjects with MDD (Fig. 1C), the duration of MDD positively correlated with densities of PNNs in CA1 SO (Supplementary Figure 1A). This is in line with preclinical evidence demonstrating that animals exposed to chronic social defeat display increased PNNs in the hippocampus ^90^. Over time, increased PNNs may alter E/I balance contributing to decreased hippocampal sharp wave ripple events that negatively affect mood and memory ^91, 92^. Increased expression of VAMP2 in subjects with MDD is in line with preclinical evidence for glucocorticoid-mediated calcium influx increases in hippocampal CA regions which perturbs plasticity, potentially resulting in poorer cognitive performance ^93^. Increased VAMP2 is also in agreement with evidence indicating altered synaptic markers in MDD ^94^. Decreases in PNNs may result in reduced stabilization of synapses and contribute to synaptic alterations, potentially resulting in memory impairment, memory generalization, and cognitive deficits observed in MDD ^95^. An extensive number of studies implicate glial cell alterations in the pathology of depression (for review see ^96^). In patients with SUD/MDD, we observed decreased expression of *Gfap*. A similar nonsignificant trend for decreased *Gfap* expression was observed in subjects with MDD. In our ANCOVA models, *Gfap* expression was significantly affected by history of SSRI exposure, which was positively associated with *Gfap* expression (Supplementary Figure 1B), suggesting that therapeutic effects of SSRIs may in part normalize *Gfap* levels. Our observed increase of *Ctss* gene expression may represent a heightened inflammatory state in the hippocampus of subjects with MDD, which has been extensively implicated in MDD ^97, 98^, including in the hippocampus in a cohort that largely overlaps with our cohort of subjects with MDD ^99^.

## TECHNICAL CONSIDERATIONS

Several factors represent challenges in examining molecular pathology in the brain of subjects with SUD, including polysubstance use, comorbidity with MDD, and pharmacological treatments. However, considering that this is representative of the real-world population of SUD it is important to examine the molecular pathology when these factors are present in the same subjects when considering development of therapeutic treatment strategies for this population. Our cohort consists of extensive information including retrospective clinical assessments, history of specific drugs of abuse and presence or absence of drugs of abuse and psychiatric medications in the blood at death from toxicology reports that can be accounted for using ANCOVA analyses, as conducted in our previous study using this same cohort ^43^. Furthermore, the inclusion of samples from rhesus monkeys with chronic alcohol use provides a valuable cause and effect comparison for the most common drug of abuse in our human subjects. The development of additional collections of NHP brain samples containing subjects self-administering specific drugs of abuse alone or in combination would further aide in interpreting human postmortem brain studies and guiding strategies for novel treatment development. For our microscopy studies, use of WFA lectin provides a standard view of a specific population of PNNs, as it binds to non-sulfated N-acetyl-D-galactosamine ^45^. There are other populations of PNNs that can be detected with alternative labeling methods which may surround complementary populations of neurons ^100^. Future studies with additional markers for PNN core proteins and sulfation motifs will provide insight into the specific components of PNNs in SUD. Further, our fresh frozen postmortem tissue has inherent technical limitations including lack of ability to conduct PVB-immunolabeling as well as reliable immunolabeling of synaptic markers, therefore mRNA measures were used for these markers. Measurements of mRNA by qPCR do not identify the cell-types responsible for synthesis or functional activity of any of the genes measured.

## CONCLUSIONS

In summary, our findings represent the first evidence for increased numerical densities of PNNs in the hippocampus of human subjects with SUD. Increased densities of hippocampal PNNs, together with decreased expression of ECM proteolytic genes and altered synaptic markers supports preclinical studies that suggest that PNNs may stabilize reward memories. Thus, PNNs and ECM molecules may represent promising targets for addressing cue-induced relapse in SUD. In comparison, subjects with MDD displayed generally opposite effects for many of the main outcome measures and subjects with SUD with a comorbid diagnosis of MDD suggesting that ECM changes in MDD oppose changes in SUD. These findings highlight the importance of considering comorbidities regarding SUD pathology and treatment strategies.

## Funding

This work was funded by support from the National Institute of Mental Health (R01 MH125833 and R01 MH117460), the National Institute of Alcoholism and Alcohol Abuse (F31 AA030166 and R01 AA029023), and the Postmortem Brain Core of the Center for Psychiatric Neuroscience, funded through an IDeA COBRE award from the National Institute of General Medical Sciences (P30 GM103328).

## Acknowledgements

The authors deeply appreciate the invaluable contributions made by the families consenting to donate brain tissue and be interviewed. We also gratefully acknowledge the support of the staff of the Cuyahoga County Medical Examiner’s Office, Cleveland, Ohio. We acknowledge the expert assistance of Drs. James C. Overholser, George Jurjus, Lisa C. Konick, and of Lesa Dieter in establishing the psychiatric diagnoses, acquiring written consent and in collecting the tissues. For some of the subjects, the services of Timothy M. De Jong in acquiring written consent and Lisa Larkin and Nicole Herbst in tissue collection are gratefully acknowledged.

## Conflict of Interest Statement

The authors have no competing financial interests to disclose.

## Data Availability Statement

The data and original contributions presented in this study are included in the manuscript or supplemental materials. Further inquiries can be directed to the corresponding author.

## Notes

### Competing Interest Statement

The authors have declared no competing interest.

### Author Declarations

The Institutional Review Boards of the University of Mississippi Medical Center, Jackson, MS and the University Hospitals Cleveland Medical Center, Cleveland, OH gave ethical approval for this work.

### Summary of Updates

The manuscript has been revised to update the figures, results section, introduction, and discussion.

